# Artificial Intelligence Model for Detecting Duodenal Endoscopic Changes on Images of Functional Dyspepsia

**DOI:** 10.1101/2022.04.26.22274340

**Authors:** Hiroshi Mihara, Sohachi Nanjo, Iori Motoo, Takayuki Ando, Haruka Fujinami, Ichiro Yasuda

## Abstract

**Introduction and Aims:** Recently, it has been suggested that the duodenum may be the pathological locus of functional dyspepsia (FD), but there are few reports on the presence of duodenal changes on imaging in FD.

**Methods:** Duodenal images acquired at our hospital with the presence or absence of the term “functional dyspepsia” on the electronic medical record, and *H. pylori* (HP) infection status on the Japan Endoscopy Database were obtained. The Google Cloud Platform AutoML Vision (single-label classification) was used to classify FD/HP current infection, versus FD/HP uninfected patients. We constructed an AI model to distinguish four groups, which included FD/HP current infection, FD/HP uninfected, non-FD/current infection, and non-FD/HP uninfected, and calculated the sensitivity, specificity, and AUC. Patient images with other organic diseases such as gastrointestinal cancer, peptic ulcer, postoperative abdominal organs, and gastroesophageal reflux disease were excluded. Narrow band imaging and dye-spread images were also excluded.

**Results:** The overall AUC of the four groups was 0.47 (FD/HP current infection 0.20, FD/HP uninfected 0.35, non-FD/current infection 0.46, non-FD/HP uninfected 0.74). Next, using the same images, we constructed a model to determine the presence or absence of FD in HP infected patients only. The sensitivity, specificity, positive predictive value, negative predictive value, and AUC were 71.4%, 66.7%, 50.0%, 83.3%, and 0.84, respectively. However, when we constructed a model to determine the presence of FD only in uninfected HP patients, the sensitivity, specificity, positive predictive value, negative predictive value, and AUC were 58.3%, 100%, 100%, 77.3%, and 0.85, respectively.

**Conclusion:** The results suggest that the duodenal imaging AL model may be able to determine the presence or absence of FD to a certain degree in HP-infected or uninfected patients.

## Introduction

The prevalence of functional dyspepsia (FD) is high, ranging from 11.0% to 23.8% in Europe, 15% in the U.S., and 7.0% to 17% in Japan[1] [2]. It has been reported that 53% patients presenting with upper abdominal symptoms have FD[3]. Regarding dyspepsia symptoms and *H. pylori* (HP) infection, it has been reported that eradication therapy improves dyspepsia symptoms in a certain percentage of HP infected patients, and a new disease entity called *H. pylori*-associated dyspepsia has been defined[4]. Recently, it was reported that *E. coli* forms endoscopically visible biofilms in the terminal ileum and ascending colon of irritable bowel syndrome (IBS) patients, and dysbiosis and microinflammation can occur even in areas without biofilm formation[5]. Although there are some cross-sectional studies indicating that antral gastritis and some endoscopic findings (linear redness in the antral region) were associated with dyspepsia symptoms, in general FD does not have distinctive features [6]. Recently, it was reported that the pathological locus of FD is in the duodenum, where dysbiosis and microinflammation occur[7].

Imaging artificial intelligence models (AI) to detect gastrointestinal endoscopic lesions have been developed, and some of them are already in clinical use[8]. A meta-analysis of the diagnostic performance for HP infection reported a sensitivity of 0.87, a specificity of 0.86, and an area under the curve (AUC) of 0.92[9], which is a higher diagnostic accuracy than that of medical practitioners.

Complex AI models such as those developed for imaging applications require deep learning algorithms and generally can only be built using Python libraries and programming expertise. However in recent years, it has become possible to build models without such programming expertise, using tools such as Google Cloud Platform (Google Inc. Mountain View, CA Available at: http://cloud.google.com/vision/. Accessed 13 Feb 2022). Normally, it is necessary for investigators to provide training datasets to the AI; however, such data do not exist for functional gastrointestinal diseases, which do not show abnormalities even on endoscopy. However, if the presence or absence of symptoms is used as training datasets to build the AI, it is thought that the AI may be able to detect minute changes in the duodenum that cannot be detected by human observers. The ultimate goal is to build an AI model that can diagnose FD and predict treatment efficacy and prognosis based on patient background and endoscopic images. In this study, we aimed to explore whether it is possible to build an image model that can identify the name of FD disease from duodenal image AI.

## Materials and Methods

### 1) Differentiation of HP current infection, uninfected, and post-eradication

We investigated if the Cloud AI construction service could produce a model to determine the status of HP infection with the same success as previously reported[9], and if the resultant AI could determine the status after eradication of the infection. We included gastric images acquired from 2020 to 2022 (Olympus, GIF-H290Z) along with the HP infection status from the Japan Endoscopy Database (JED) in our hospital. We then built an AI model to distinguish HP current infection, uninfected, and post-eradication, as well as current infection versus uninfected on the stomach images using Google Cloud Platform AutoML Vision (single label classification). Ulcer lesions, food residue, post-gastrectomy, and narrow band, magnification images were excluded. This entire process was performed by one physician (HM) and one medical technician (K).

### 2) Differentiation of duodenal images in functional dyspepsia by HP infection status

We investigated whether the cloud AI tool could be used to build a model that could distinguish between the duodenal images of patients with FD disease names recorded for insurance purposes in their electronic medical records versus asymptomatic patients.

We acquired duodenal endoscopic images of FD patients with the disease name in the medical record and patients with abnormal health examinations whose endoscopic report showed no abnormalities from January 2018 to December 2020 and built an AI model to distinguish the four groups of FD and HP infection status (FD/HP current infection (+), FD/HP uninfected(-), non-FD/HP(+), and non-FD/HP(-)), as well as each two groups (FD, non-FD among HP(+) or HP(-)). For this part, the post-eradication patients were excluded. Most of the patients with abnormal physical examinations are those who were found to have abnormalities during barium screening for gastric cancer. The HP detection model was constructed similarly. Since it is impossible to determine whether the amount of air delivered differs or whether the duodenum is difficult to extend despite adequate air delivery is itself an original finding, to reduce selection bias and maintain accuracy for eventual use in actual clinical practice, we employed as many images as possible that had been captured and used them as training data without excluding images with insufficient extension.

### Artificial neural network (ANN) programming and training

Using the Google Cloud AutoML Vision platform, the training, validation, and the test images were randomly selected from the dataset automatically and were all independent of each other. We used all of the image datasets (training images 80%, validation images 10% and the test images 10%) to build the AI model. The number of images selected for the test set was proportional to the number in the training set. The algorithm was naïve to the images in the training set. Sixteen nodes (2 h) were used to train the algorithm on the AutoML Vision cloud-based graphical processing units. A single label image classification architecture was utilized.

### Statistical analysis

The code for Google Cloud AutoML Vision platform has not been made publicly available by Google, the company responsible for its development. The larger the sample size for image model building, the better, but there is no way to calculate the minimum number, and Google AutoML requires 100 images for each label to build. Since increasing the sample size did not increase the accuracy any further, it was determined that this sample size was the optimal sample size for this project.

AutoML Vision provided metrics that are commonly used by the AI community. These include the precision (positive predictive values) and recall (sensitivity) for a stated threshold and the area under the curve (AUC). We also provided confusion matrices for each model, which cross-referenced the true labels against those predicted by the deep learning mode[10].

Using extracted binary diagnostic accuracy data, we constructed contingency tables showing the calculated specificity at a threshold of 0.5. The contingency tables showed true-positive, false-positive, true-negative, and false-negative results.

## Results

### (1) HP infection model of the stomach

We used 2863 images from 81 currently infected patients, 3196 images from 98 uninfected patients, and 3436 images from 105 post-eradication patients. Images with blurred focus and high gastric juice content were excluded, and approximately 30 images per patient were used. The model was constructed within 1.2 hours and the cost was approximately 30 U.S. dollars.

The overall AUC of the three groups was 0.63 (Table 1). It was difficult to differentiate eradicated stomachs from infected and uninfected stomachs.

**Table 1.** HP infection model of the stomach; AUC and the contingency tables

|  | AUC |  |  |
| --- | --- | --- | --- |
| All labels | 0.64 |  |  |
| HP current infection | 0.62 |  |  |
| Post eradication | 0.65 |  |  |
| Uninfected | 0.72 |  |  |
|  | Predictive label |  |  |
| True label | HP current infection | Post eradication | Uninfected |
| HP current infection | 40% | 50% | 11% |
| Post eradication | 13% | 76% | 11% |
| Uninfected | 10% | 43% | 47% |

We next constructed a model to distinguish between currently infected and uninfected stomachs using the same image set, and found a sensitivity of 0.76, a specificity of 0.74, and an AUC of 0.85 for currently infected stomachs (Table 2). The average precision (positive predictive value) of the algorithm was 77.0% and the recall was 77.8% based on the automated training and testing by the Google Cloud AutoML Vision in Fig. 1. The precision recall curves were generated for each individual label as well as for the algorithm overall. We adopted a threshold value of 0.5 to yield a balanced precision and recall. When comparing the duodenal images of patients with FD, we decided to exclude, from the outset, patients who had undergone eradication.

**Table 2.** HP infection model of the stomach; AUC and the contingency tables

|  | AUC |  |
| --- | --- | --- |
| All labels | 0.84 |  |
| HP current infection | 0.81 |  |
| Uninfected | 0.86 |  |
|  | Predictive label |  |
| True label | HP current infection | Uninfected |
| HP current infection | 75% | 25% |
| Uninfected | 21% | 79% |

**Fig. 1.**
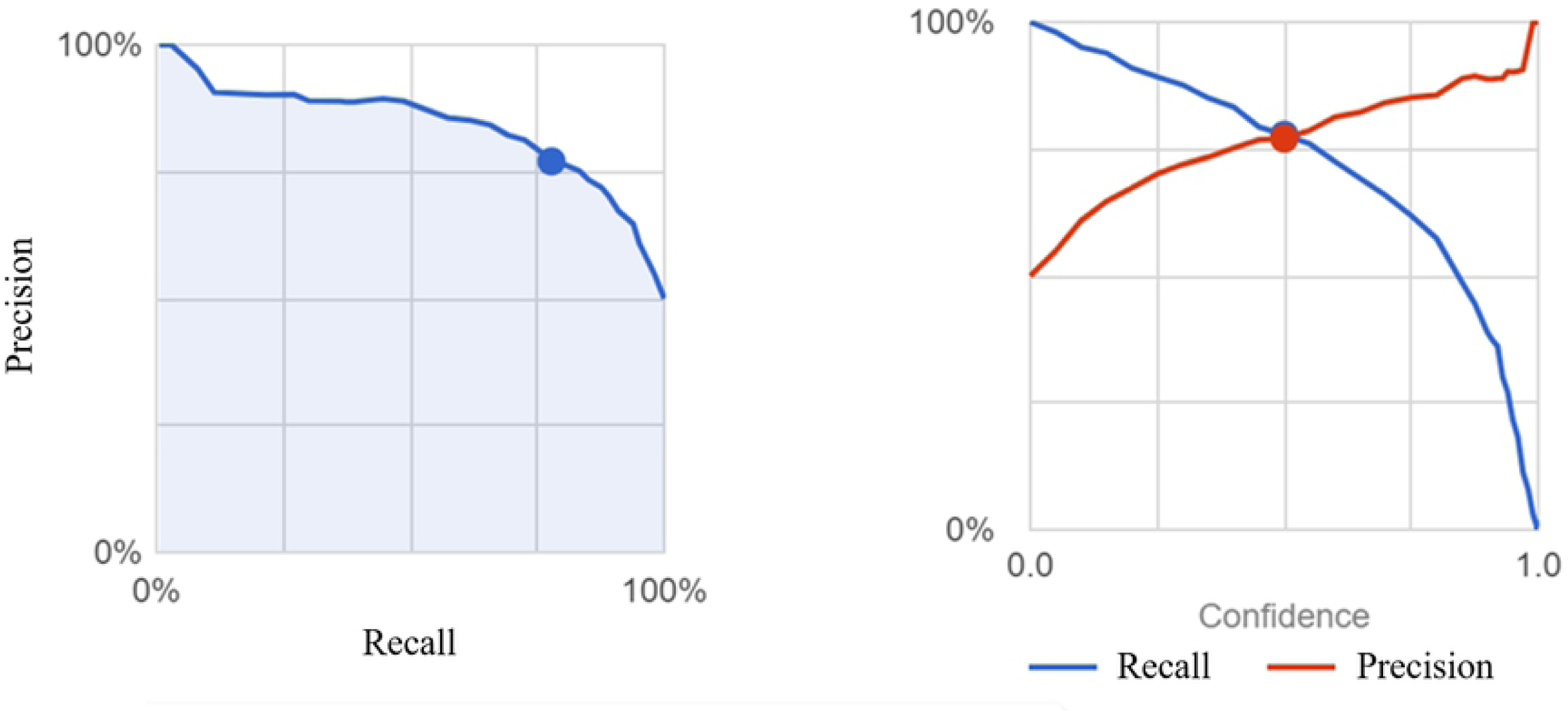
The positive predictive values, recall, and area under the curve (AUC) for a threshold value of 0.5 in HP infected stomachs are shown.

### (2) FD models

The dataset included 70 images from 32 FD/HP patients with current infection, 121 images from 35 FD/HP uninfected patients, 147 images from 39 patients with non-FD/HP current infection and 156 images from 33 non-FD/HP uninfected patients. We used less than 10 images per patient, and they included the region from the duodenal bulbus to the descending portion of the duodenum.

The model was constructed within 2 hours and cost approximately 30 U.S. dollars. The overall AUC of the four groups was 0.46 (Table 3). Since the AUCs of the FD/HP(+) cases were the lowest and those of patients with non-FD/HP(-) were the highest, it was thought that the presence or absence of HP infection affected the accuracy of the FD discrimination. Therefore, the same data set was reused as input for a model to differentiate only the presence or absence of FD with or without HP infection (Table 4). The sensitivity, specificity, positive and negative predictive values, and the AUC of the model to determine the presence or absence of FD in currently infected HP patients were 71.4%, 66.7%, 50.0%, 83.3%, and 0.84, respectively. In uninfected HP patients they were 58.3%, 100%, 100%, 77.3%, and 0.85, respectively. The precision and recall of the algorithm were 68.2% and 68.2% in HP(+) and 82.76% and 82.76% in HP(-), respectively (Figure 2).

**Table 3.** FD and HP infection model of the duodenum; AUC and the contingency tables AUC

| True label | FD/HP(-) | FD/HP(+) | Non-FD/HP(-) | Non-FD/HP(+) |
| --- | --- | --- | --- | --- |
| FD/HP(-) | 50% | 25% | - | 25% |
| FD/HP(+) | 14% | 14% | 14% | 57% |
| Non-FD/HP(-) | 12% | 6% | 59% | 24% |
| Non-FD/HP(+) | 44% | 13% | 6% | 38% |

**Table 4.**
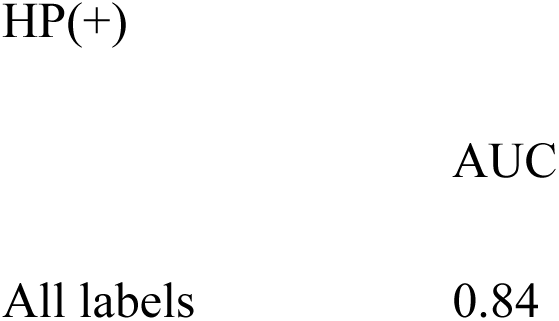
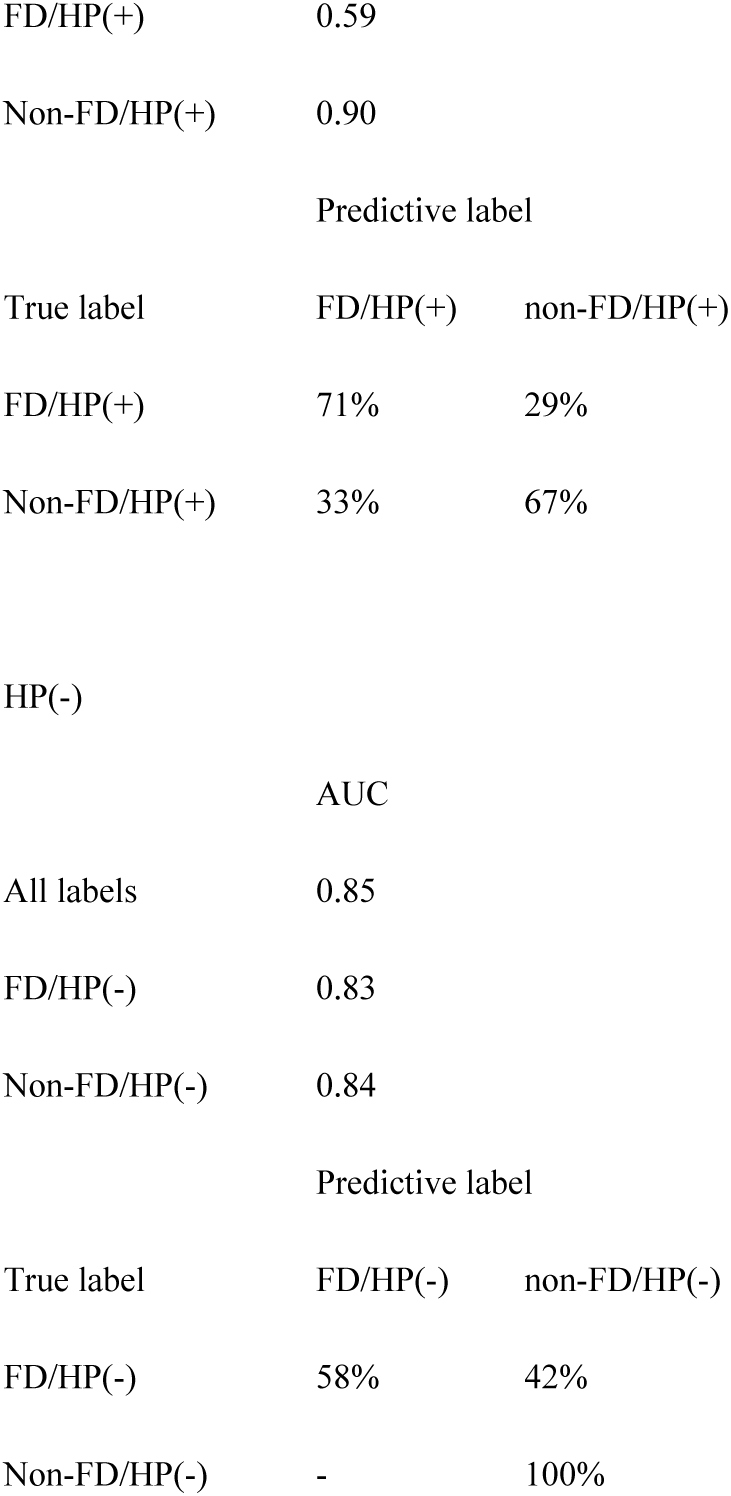
FD model of the duodenum depending on HP status; AUC and the contingency tables

**Fig. 2.**
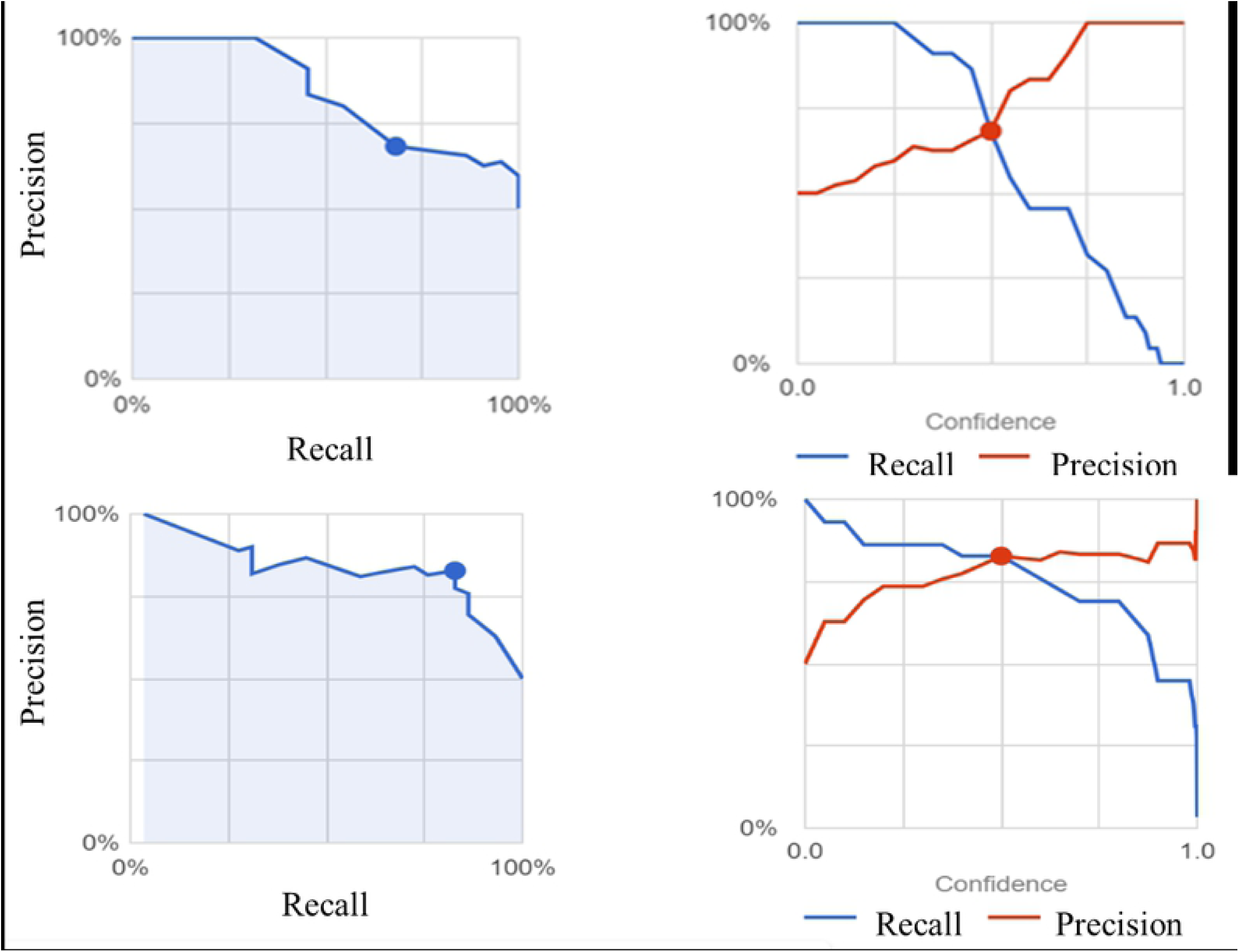
The positive predictive values, recall, and area under the curve (AUC) for a threshold value of 0.5 in FD dependent on HP infection status are shown. A;HP(+), B;HP(-).

Example images with high FD scores (Figure 3) and high normal scores (Figure 4) in HP infected patients are indicated.

**Fig. 3.**
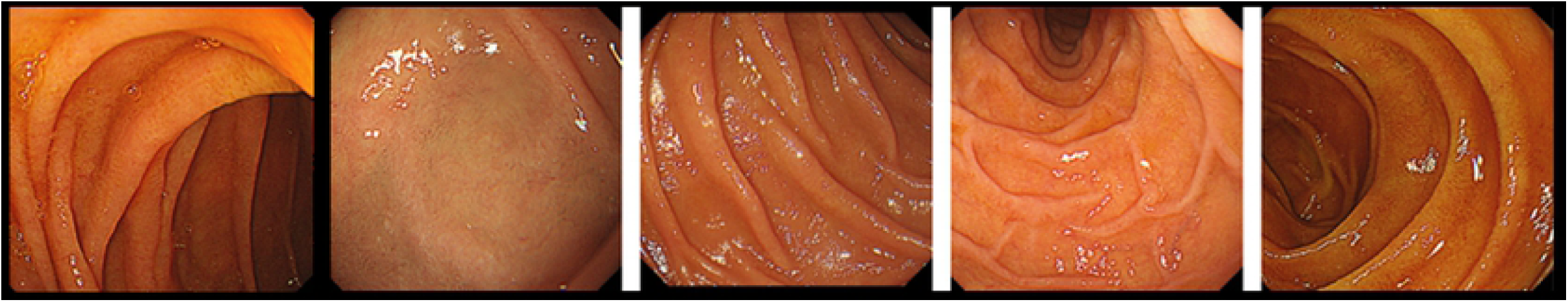
Images that scored high (0 to 1) in the duodenal image AI model for detecting the <u>absence of FD</u> constructed only in HP-infected patients are shown.

**Fig. 4.**
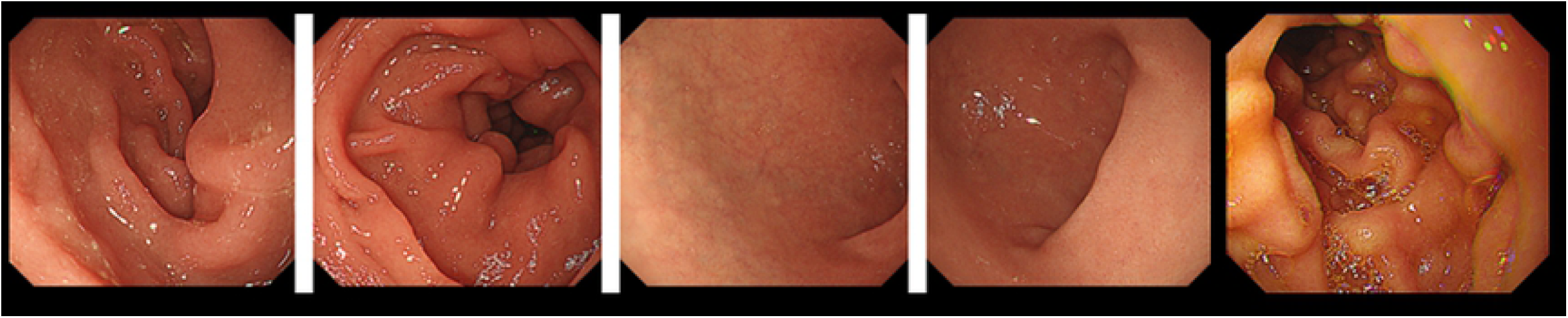
Images that scored high (0 to 1) in the duodenal image AI model for detecting the <u>presence of FD</u> constructed only in HP-infected patients are shown.

## Discussion/Conclusion

We developed an AI model to distinguish between FD and HP infection on duodenal images. It was relatively easy for the AI to distinguish between current HP infection versus uninfected, but difficult to distinguish those versus post-eradication. To date, no imaging model has been reported that differentiates post-eradication stomachs from others, and this report is considered the first finding. The AI model was able to distinguish between FD and HP infection. This is a novel finding and may lead to an endoscopic diagnosis of functional gastrointestinal diseases, as the duodenum of FD patients is usually considered normal, but there are minute differences, which can be detected by deep learning algorithms. The results also suggest that *H. pylori* infection of the stomach causes changes in the endoscopic image of the duodenum. From the pathophysiology, it is assumed that the AI can detect the mild edema caused by microinflammation. However the limitation of deep learning methods is that ultimately, it is impossible to know which image feature is being used by the AI for the classification. Other limitations of this study include 1) the FD was classified by the presence of disease names for insurance rather than Rome criteria, 2) the population was prescribed acotiamide, 3) many of the cases were considered to have postprandial complaints syndrome, and 4) background factors such as age and gender were not taken into account. In addition, the HP infection data was based on the JED, and test results such as urinary antibodies and fecal antigens were not always confirmed. Therefore, the reliability and reproducibility of the model need to be further analyzed in an external validation cohort of actual clinical cases.

The pathogenesis of FD is multifactorial, and in addition to microinflammation of the duodenum, gastrointestinal motility, sensory, mucosal integrity, and dysregulation of the gut-brain axis are involved. The ultimate goal is to construct a model to predict treatment response and prognosis based not only on basic background factors such as age, gender, BMI, smoking, and alcohol consumption, *H*.*pylori* eradication status, but also on the degree of atrophy, history of drug administration such as acotiamide, PPIs, and NSAIDs, comorbidities such as DM, and FD values obtained from duodenal imaging AI such as those constructed here, which will be of great clinical benefit. On the other hand, a dataset of at least 1000 cases is required to construct a prediction model, and in this study, we did not intentionally collect patient background information because the purpose was only to construct a duodenal image AI.

In general, it takes an engineer who is an expert in deep learning to manually create AI models with algorithms specific to each study. However, the code-free deep learning approach adopted in this study has the potential to improve clinicians’ access to deep learning tools including prediction models [11]. Other research groups have reported automated deep learning approaches, which were developed without coding experience for medical image classification and otoscopic diagnosis[10, 12, 13]. The accuracy is expected to vary with different endoscope light source settings and scopes. Other than the option of using a very large dataset from a large number of facilities as training data, there is also an option to build a simple AI for each facility individually. To our knowledge, this is the first report of a meaningful AI model, which was built using the presence or absence of symptoms as training data. In the future, differences in diseases, which may have no apparent abnormalities may be distinguished by AI models. As described above, we created an AI model based on duodenoscopy images and HP infection status using FD symptoms as training data without actual coding expertise. It is expected that an AI model with symptoms included in the training data may be able to differentiate endoscopic images of functional gastrointestinal diseases from asymptomatic patients.

## Data Availability

Data Availability Statement Data cannot be shared for confidentiality reasons. Queries about the data should be directed to the corresponding author.

## Acknowledgements

We thank Ayaka Maeda, Masaya Hiraki, Shun Kuraishi, and Kenji Ogawa, medical engineering technicians at the Medical Device Management Center, University of Toyama Hospital, for their support in collecting and organizing the images.

The summary of this study was presented at the 18th Annual Meeting of the Japanese Gastroenterological Association Core Symposium 4: Pathogenesis and Treatment of Functional Gastrointestinal Diseases [Basic and Clinical Frontiers of Functional Gastrointestinal Diseases].

## Statement of Ethics

The study protocol was approved by the Ethics Committee of the Toyama University hospital (approval No. R2021032). All methods were performed in accordance with the relevant guidelines and regulations as well as with the Declaration of Helsinki. The study design was accepted by the ethics committee on the condition that a document declaring an opt-out policy by which any potential patients and/or their relatives could refuse to be included, was uploaded to the website of the Toyama University hospital.

## Author Contributions

All authors contributed to the conception and design of the study. Hiroshi Mihara collected images and Sohachi Nanjo, Iori Motoo, Takayuki Ando and Haruka Fujinami reviewed the images. Hiroshi Mihara provided training in automated deep learning models. Hiroshi Mihara drafted the manuscript and all authors confirmed the final version of manuscript.

## Data Availability Statement

Data cannot be shared for confidentiality reasons. Queries about the data should be directed to the corresponding author.

## References

1. Mahadeva S, Goh KL. Epidemiology of functional dyspepsia: a global perspective. World J Gastroenterol. 2006;12(17):2661–6. doi: 10.3748/wjg.v12.i17.2661. PubMed PMID: 16718749; PubMed Central PMCID: PMCPMC4130971.

2. Oshima T, Miwa H. Epidemiology of Functional Gastrointestinal Disorders in Japan and in the World. J Neurogastroenterol Motil. 2015;21(3):320–9. doi: 10.5056/jnm14165. PubMed PMID: 26095436; PubMed Central PMCID: PMCPMC4496905.

3. Okumura T, Tanno S, Ohhira M, Tanno S. Prevalence of functional dyspepsia in an outpatient clinic with primary care physicians in Japan. J Gastroenterol. 2010;45(2):187–94. doi: 10.1007/s00535-009-0168-x. PubMed PMID: 19997854.

4. Stanghellini V, Chan FK, Hasler WL, Malagelada JR, Suzuki H, Tack J, et al. Gastroduodenal Disorders. Gastroenterology. 2016;150(6):1380–92. doi: 10.1053/j.gastro.2016.02.011. PubMed PMID: 27147122.

5. Baumgartner M, Lang M, Holley H, Crepaz D, Hausmann B, Pjevac P, et al. Mucosal Biofilms Are an Endoscopic Feature of Irritable Bowel Syndrome and Ulcerative Colitis. Gastroenterology. 2021;161(4):1245–56 e20. doi: 10.1053/j.gastro.2021.06.024. PubMed PMID: 34146566; PubMed Central PMCID: PMCPMC8527885.

6. Tahara T, Arisawa T, Shibata T, Nakamura M, Okubo M, Yoshioka D, et al. Association of endoscopic appearances with dyspeptic symptoms. J Gastroenterol. 2008;43(3):208–15. doi: 10.1007/s00535-007-2149-2. PubMed PMID: 18373163.

7. Miwa H, Oshima T, Tomita T, Fukui H, Kondo T, Yamasaki T, et al. Recent understanding of the pathophysiology of functional dyspepsia: role of the duodenum as the pathogenic center. J Gastroenterol. 2019;54(4):305–11. doi: 10.1007/s00535-019-01550-4. PubMed PMID: 30767076; PubMed Central PMCID: PMCPMC6437122.

8. Kudo SE, Misawa M, Mori Y, Hotta K, Ohtsuka K, Ikematsu H, et al. Artificial Intelligence-assisted System Improves Endoscopic Identification of Colorectal Neoplasms. Clin Gastroenterol Hepatol. 2020;18(8):1874–81 e2. doi: 10.1016/j.cgh.2019.09.009. PubMed PMID: 31525512.

9. Bang CS, Lee JJ, Baik GH. Artificial Intelligence for the Prediction of Helicobacter Pylori Infection in Endoscopic Images: Systematic Review and Meta-Analysis Of Diagnostic Test Accuracy. J Med Internet Res. 2020;22(9):e21983. doi: 10.2196/21983. PubMed PMID: 32936088; PubMed Central PMCID: PMCPMC7527948.

10. Faes L, Wagner SK, Fu DJ, Liu X, Korot E, Ledsam JR, et al. Automated deep learning design for medical image classification by health-care professionals with no coding experience: a feasibility study. Lancet Digit Health. 2019;1(5):e232–e42. doi: 10.1016/S2589-7500(19)30108-6. PubMed PMID: 33323271.

11. Korot Eea. Code-free deep learning for multi-modality medical image classification.. Nat Mach Intell 2021.

12. Livingstone D, Chau J. Otoscopic diagnosis using computer vision: An automated machine learning approach. Laryngoscope. 2020;130(6):1408–13. doi: 10.1002/lary.28292. PubMed PMID: 31532858.

13. Ito Y, Unagami M, Yamabe F, Mitsui Y, Nakajima K, Nagao K, et al. A method for utilizing automated machine learning for histopathological classification of testis based on Johnsen scores. Sci Rep. 2021;11(1):9962. doi: 10.1038/s41598-021-89369-z. PubMed PMID: 33967273; PubMed Central PMCID: PMCPMC8107178.

